# Highly pathogenic coronavirus N protein aggravates lung injury by MASP-2-mediated complement over-activation

**DOI:** 10.1101/2020.03.29.20041962

**Authors:** Ting Gao, Mingdong Hu, Xiaopeng Zhang, Hongzhen Li, Lin Zhu, Hainan Liu, Qincai Dong, Zhang Zhang, Zhongyi Wang, Yong Hu, Yangbo Fu, Yanwen Jin, Kaitong Li, Songtao Zhao, Yongjiu Xiao, Shuping Luo, Lufeng Li, Lingfang Zhao, Junli Liu, Huailong Zhao, Yue Liu, Weihong Yang, Jing Peng, Xiaoyu Chen, Ping Li, Yaoning Liu, Yonghong Xie, Jibo Song, Lu Zhang, Qingjun Ma, Xiuwu Bian, Wei Chen, Xuan Liu, Qing Mao, Cheng Cao

## Abstract

An excessive immune response contributes to SARS-CoV, MERS-CoV and SARS-CoV-2 pathogenesis and lethality, but the mechanism remains unclear. In this study, the N proteins of SARS-CoV, MERS-CoV and SARS-CoV-2 were found to bind to MASP-2, the key serine protease in the lectin pathway of complement activation, resulting in aberrant complement activation and aggravated inflammatory lung injury. Either blocking the N protein:MASP-2 interaction or suppressing complement activation can significantly alleviate N protein-induced complement hyper-activation and lung injury *in vitro* and *in vivo*. Complement hyper-activation was also observed in COVID-19 patients, and a promising suppressive effect was observed when the deteriorating patients were treated with anti-C5a monoclonal antibody. Complement suppression may represent a common therapeutic approach for pneumonia induced by these highly pathogenic coronaviruses.

**One Sentence Summary:** The lectin pathway of complement activation is a promising target for the treatment of highly pathogenic coronavirus induced pneumonia.

## Main Text

Severe acute respiratory syndrome (SARS), that was initially reported in Guangdong, China, in November 2002, is a highly contagious and deadly respiratory disease (*1, 2*). Severe acute respiratory syndrome coronavirus (SARS-CoV) was identified as the novel etiological agent of this disease. Nearly a decade after the SARS outbreak, a new zoonotic coronavirus, Middle East respiratory syndrome (MERS) coronavirus (MERS-CoV), was identified as the etiological agent of MERS (*3*). Recently, a new coronavirus, SARS-CoV-2 was first identified in Wuhan, China and spread rapidly to other provinces in china and all over the world. As of 19 March 2020, SARS-CoV-2 has infected more than 230000 people with a fatality rate of 4.2% (https://www.who.int/emergencies/diseases/novel-coronavirus-2019/situation-reports/). Infection of the virus caused severe atypical pneumonia similar to SARS-CoV infection (*4*). Although the pathogenesis of these diseases are being intensively investigated, it is still not well understood why the viral infections lead to respiratory failure with a high fatality rate (*5*). The SARS-CoV nucleocapsid (N) protein is a 46-kDa viral RNA-binding protein sharing only 20-30% homology with the N proteins of other known coronaviruses(*6*), whereas N proteins of the highly pathogenic coronaviruses are more similar, including SARS-CoV-2 (91%) and MERS-CoV (51%) by BLASTP (*5, 7, 8*). The N protein is one of the most abundant viral structural proteins in patient sera during SARS-CoV infection (*9*). Potentially N protein plays a role in the virus pathogenesis as the pre-administration of N protein, but not other viral proteins, via recombinant vaccinia virus or Venezuelan equine encephalitis virus replicon particles resulted in severe pneumonia in aged mice challenged with SARS-CoV (*10, 11*).

The complement system functions as an immune surveillance system that rapidly responds to infection. Activation of the complement system resulted in pathogen elimination and acute or chronic inflammation. Nevertheless, dysregulated complement activation has been implicated in the development of acute lung diseases induced by highly pathogenic viruses (*12, 13*). The complement system can be activated via the classical pathway (CP), the lectin pathway (LP), or the alternative pathway (AP) (*14*). In the LP, mannan-binding lectin (MBL) (or ficolins) binds to carbohydrate arrays of mannan and N-acetylglucosamine residues on the surfaces of the viruses or the surfaces of virus-infected cells, resulting in the activation of MBL-associated serine protease-2 (MASP-2), the only known MBL-associated protease that can directly initiate the complement cascade (*15, 16*). MBL binds to SARS-CoV-infected cells in a dose-dependent, calcium-dependent, and mannan-inhibitable manner *in vitro*, enhancing the deposition of complement C4 on SARS-CoV (*17*). The N-linked glycosylation site N330 on the SARS-CoV spike (S) protein is critical for the specific interactions with MBL (*18*). Although higher levels of activated complement C3 and C4 fragments were found in SARS patients, indicating the activation of complement pathways (*19, 20*), the mechanism of SARS-CoV-induced complement activation is not well understood. It is also unknown whether a similar pathogenesis occurs in SARS-CoV-2 infection.

Previously, we demonstrated that the SARS-CoV N protein interacts with a number of host proteins, including MAP19 (*21*), an alternative splicing product of MASP-2. Thus, in this study, the interactions of the SARS-CoV, MERS-CoV and SARS-CoV-2 N proteins with MASP-2 were intensively investigated, and the pathological effects of these interactions on host immunity and inflammation were also elucidated, which provides a mechanism support for immunomodulation-based therapy against current COVID-19 epidemic.

## Results

### N proteins of SARS-CoV, SARS-CoV-2 and MERS-CoV interact with MASP-2

To investigate the binding between the N protein of SARS-CoV and MASP-2, and to delineate the interacting domains of the two proteins, lysates of human 293T cells expressing Flag-tagged full-length MASP-2, the N-terminal CUB1-EGF-CUB2 region, or the C-terminal CCP1-CCP1-SP region (Fig. S1A) were subjected to anti-Flag immunoprecipitation using anti-Flag antibody-conjugated agarose beads. The immunoprecipitates were next incubated with 293T cell lysates expressing GFP-tagged SARS-CoV N protein (GFP-SARS N) or truncated mutants in the presence of 2 mM CaCl_2_ or 1 mM EDTA. The adsorbates were probed with anti-Flag or anti-GFP antibodies by immunoblotting. Associations between Flag-MASP-2 and GFP-SARS N were observed only in the presence of CaCl_2_ (Fig. 1A), in agreement with the requirement of Ca^2+^ for MASP-2-MBL binding and MASP-2 auto-activation. The CCP1-CCP2-SP region of MASP-2 (Fig. 1A) and the N-terminal domain (residues 1-175) of the N protein (Fig. S1A and Fig. S1B) were crucial for the association, whereas negative controls, or other truncated regions did not bind, and GFP-tagged full-length or truncated N protein did not co-immunoprecipitate with mouse IgG conjugated beads (Fig. 1A and Fig. S1B).

**Fig 1.**
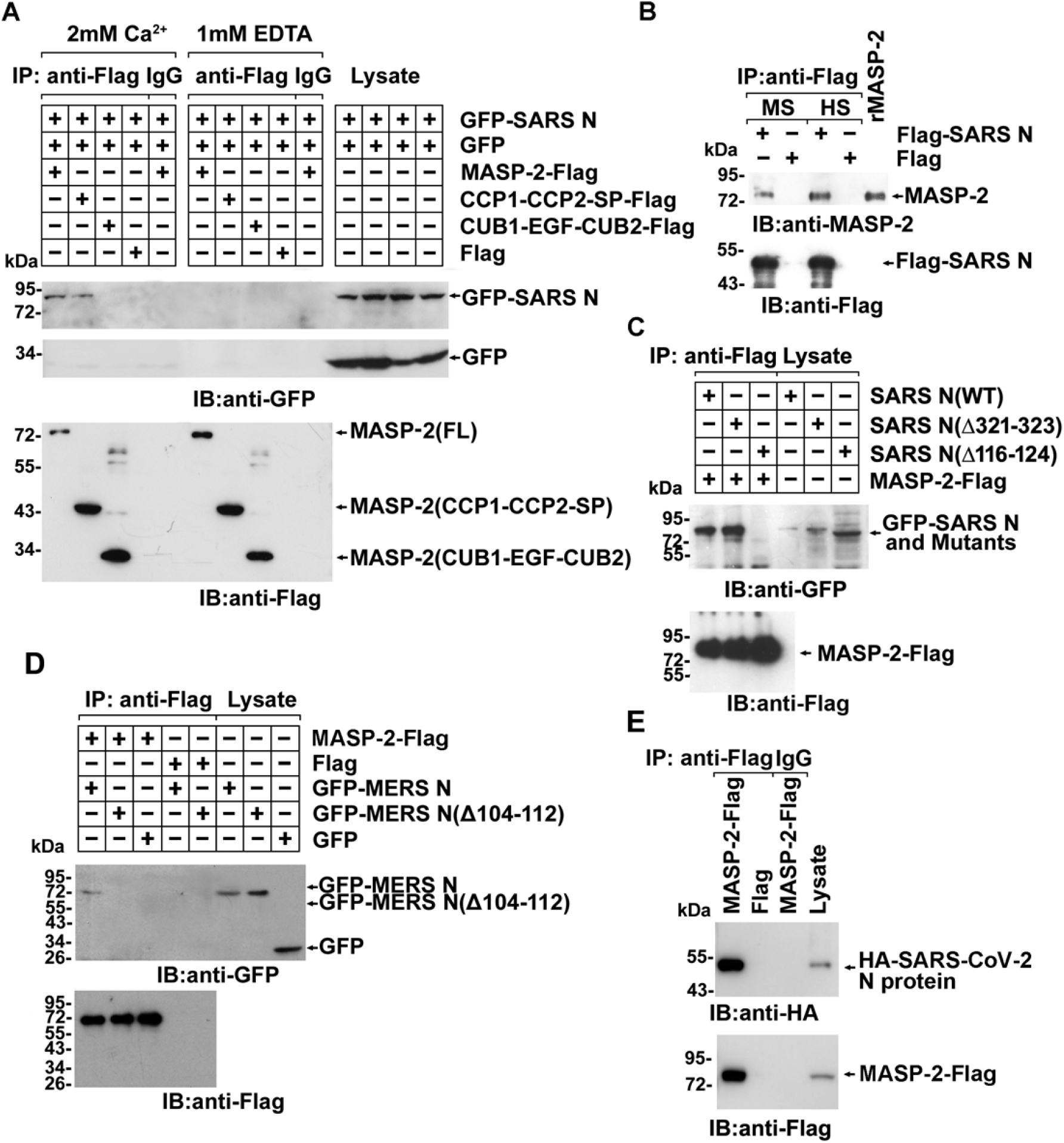
The N proteins of SARS-CoV, MERS-CoV and SARS-CoV-2 bind to MASP-2. (A) Lysates of 293T cells transfected with indicated plasmids were subjected to anti-Flag immunoprecipitation in the presence of 2 mM CaCl_2_ or 1 mM EDTA, and analyzed by immunoblotting. IgG immnuoprecipitates were used as a negative control. (B) Lysates of 293T cells transfected with indicated plasmids were mixed with human serum (HS) and mouse serum (MS) and subjected to immunoprecipitation with anti-Flag agarose beads. The absorbates were probed with indicated antibodies. Purified recombinant MASP-2 was loaded as a marker. (C)-(E) Lysates of 293T cells transfected with indicated plasmids were subjected to anti-Flag immunoprecipitation in the presence of 2 mM CaCl_2_ and analyzed by immunoblotting.

SARS-CoV N protein can be detected in patient serum as early as 1 day after the onset of symptoms (*22*). To simulate the SARS-CoV N protein associations in serum, Flag-tagged N protein (1 ng/ml) was added to human or mouse serum and precipitated with anti-Flag antibody conjugated agarose beads. The SARS-CoV N protein interacted with MASP-2 derived from both human and mouse serum (Fig. 1B). Further truncation and deletion analysis showed that amino acid residues 116-124, which are located in a coil motif of the SARS-CoV N protein (residues 115-130), were indispensable for the interaction with MASP-2 (Fig. 1C). However, the association of MASP-2 with SARS-CoV NΔ321-323, a mutant that fails to form the N protein dimer (Fig. S1C), was not much affected (Fig. 1C).

The crucial motif in SARS-CoV N protein for MASP2 interaction (residues 116-124) share a high identity with the corresponding motif in SARS-CoV-2 N (115-123) and MERS-CoV N (104-112) (Fig. S1D), suggesting that the N protein of SARS-CoV-2 and MERS-CoV will also interact with MASP-2. As expected, exogenously expressed MERS-CoV N and SARS-CoV-2 N both associated with MASP-2 (Fig. 1D and 1E), and the Δ104-112 deletion mutant of MERS-CoV N exhibited the predicted reduced association (Fig. 1D). Therefore, a common motif across coronavirus N proteins is important for MASP-2 binding.

### N proteins of SARS-CoV, MERS-CoV and SARS-CoV-2 potentiate MASP-2-dependent complement activation

The CCP1-CCP2-SP domains of MASP-2 are responsible for self-activation and substrate binding activity, which in turn mediates complement lectin pathway activation (*23*). The MASP-2: N protein association demonstrated above suggests that SARS-CoV N protein may regulate MASP-2 dimerization, activation and cleavage. To demonstrate MASP-2 dimerization, MASP-2-Flag-conjugated beads were incubated with lysates of 293T cells expressing MASP-2-Myc in the presence/absence of N protein and MBL. Binding of MASP-2-Myc to MASP-2-Flag was potentiated by the SARS-CoV N protein, at an approximately equal molar stoichiometry (Fig. 2A). Next, purified MASP-2-Flag was incubated with mannan and MBL with or without SARS-CoV N protein. Higher levels of the cleaved MASP-2 fragments (residues 445-686) resulting from MASP-2 auto-activation were produced in the presence of SARS-CoV N protein (Fig. 2B).

**Fig 2.**
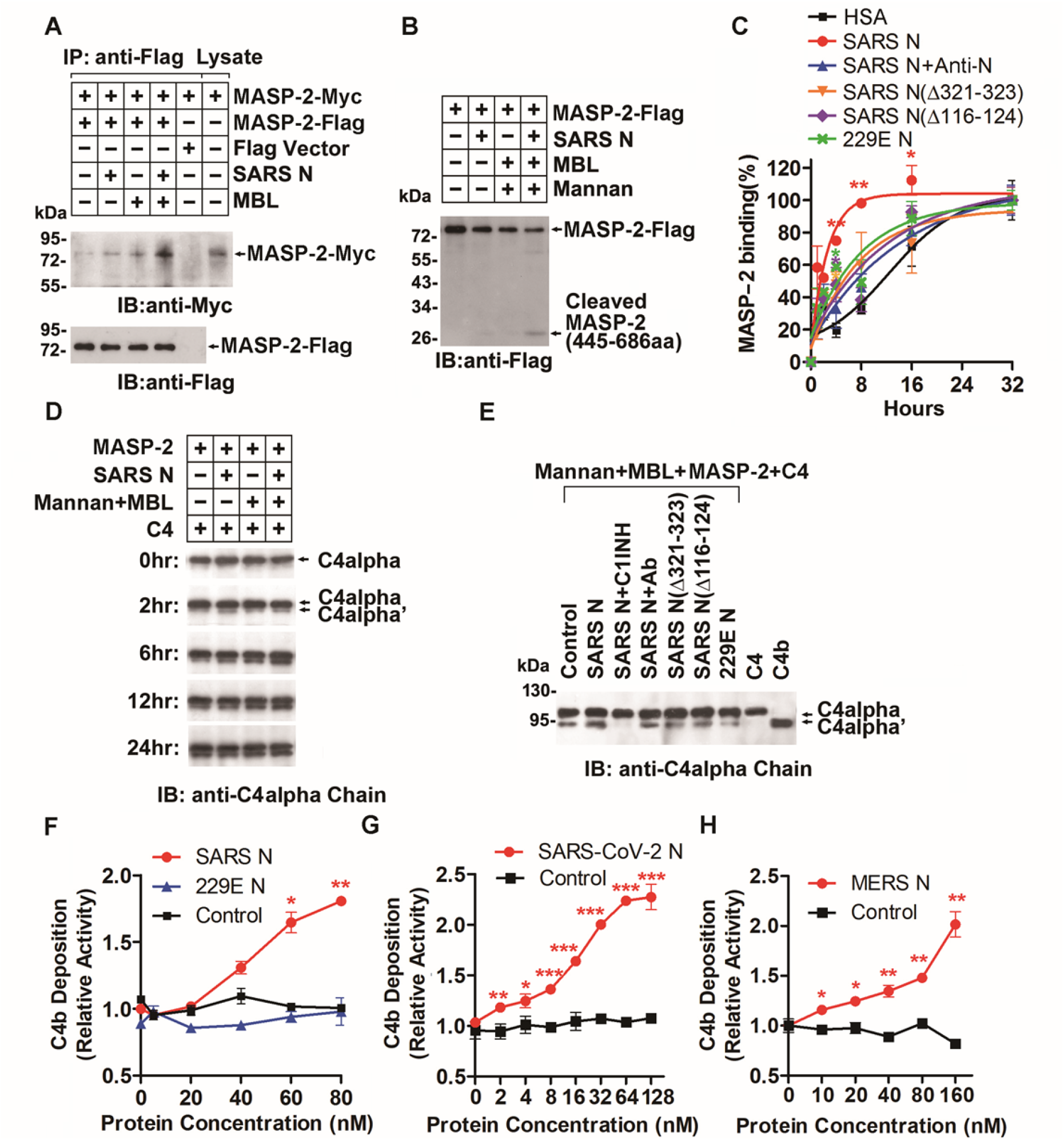
The N proteins induce MASP-2 auto-activation and C4 cleavage. (A) Lysates from cells expressing indicated proteins were mixed with purified N or MBL and subjected to anti-Flag immunoprecipitation and analyzed by immunoblotting. (B) Purified MASP-2-Flag was incubated with/without N, MBL, and mannan at 37°C for 12 hr. Cleaved MASP-2 was probed with an anti-Flag antibody. (C) Purified MASP-2 and N proteins with/without anti-N monoclonal antibody were incubated with pre-conjugated MBL in mannan-coated plates at 4°C. Binding of MASP-2 was detected with an anti-MASP-2 antibody. (D) C4 was incubated with indicated components at 37°C for 0, 2, 6, 12 and 24 hr. C4 and a cleaved C4alpha’ fragment were detected with anti-C4alpha chain antibody. (E) C4 was incubated with indicated components at 37°C for 1 hr. C4 cleavage was detected as mentioned above. Ab, anti-MASP-2 antibody. (F)-(H) C4b deposition in relation to the concentration of N proteins of SARS-CoV, HCoV-229E, SARS-CoV-2, and MERS-CoV.

To investigate the effect of SARS-CoV N protein on the MBL-binding capability of MASP-2, purified MBL and MASP-2 (*24*) were incubated in mannan-coated microplate wells at 4°C to avoid MASP-2 activation, and the dynamics of MASP-2: MBL binding were assessed using an anti-MASP2 antibody. Compared with human serum albumin (HSA), a high-concentration component in binding buffer used as a negative control for N protein, the binding of MASP-2 to MBL was significantly enhanced in the presence of SARS-CoV N protein at a relatively low concentration (1%∼0.1% of MASP-2) and Ca^2+^, and the potentiation was effectively abrogated by anti-N protein antibody (Fig. 2C). Moreover, SARS-CoV N protein bearing the Δ116-124 or Δ321-323 deletion or N protein from the less pathogenic human coronavirus 229E-CoV showed little or no effect on MASP-2:MBL binding compared with the full-length SARS-CoV N protein (Fig. 2C). These results indicated that SARS-CoV N protein-potentiated MASP-2 activation is dependent not only on MASP-2 association but also on N protein dimerization.

MASP-2 cleaves complement components C4 and C2 to generate C3 convertase in the lectin pathway of the complement system upon activation. Next, the effect of the SARS-CoV N protein on LP complement activation was assessed by C4 cleavage. Purified C4 was incubated with MASP-2, Mannan, and MBL in the presence/absence of equal molar N protein, and C4 cleavage was observed to be significantly potentiated by the SARS-CoV N protein in the presence of mannan and MBL (Fig. 2D and Fig. S2A). Accordingly, mutants of SARS-CoV N protein (Δ116-124 and Δ321-323) as well as N protein from H229E-CoV failed to promote MASP-2-mediated C4 hydrolysis (Fig. 2E and Fig. S2B). Notably, an anti-MASP-2 monoclonal antibody or C1INH, an inhibitor of MASP-2 (*25*), blocked SARS-CoV-potentiated C4 hydrolysis, suggesting that N protein-potentiated C4 cleavage was dependent on MASP-2 activation (Fig. 2E and Fig. S2B). Moreover MERS-CoV N was also found to potentiate C4 cleavage (Fig. S2C). These results indicated that N protein prompts C4 cleavage and therefore complement activation by MASP-2 association and activation.

The impact of SARS-CoV N protein on complement activation via the lectin pathway was further investigated by complement deposition assays. Purified C4 was incubated with immobilized MBL-MASP-2 complex in the presence of indicated N protein. The N protein of SARS-CoV, MERS-CoV and SARS-CoV-2 but not the H229E-CoV N potentiated C4b deposition, which was dependent on the activity of MASP-2, in a dose-dependent manner (Fig. 2F-2H). Then, immobilized mannan was incubated with C1q-depleted serum (to eliminate the classical pathway) (*26*) in the presence/absence of SARS-CoV N protein, and the deposited C3 fragments (C3b, iC3b and C3dg) were detected by an anti-activated C3 antibody. In concert with C4b deposition, the deposition of activated C3 was evidently increased along with the increase of SARS-CoV N protein levels up to ∼40 nM (Fig. S2D), suggesting an enhanced activity of C3 convertase. In addition, SARS-CoV N protein had little or no effect on activated C3 deposition in calcium-free buffer containing EGTA, which suggests that SARS-CoV N protein-potentiated C3 activation occurs through the LP but not the AP pathway, in which C3 activation is Ca^2+^-independent (Fig. S2E). As shown in Figure S2D and S2E, C3b deposition was decreased in the presence of a high concentration of N protein, possibly due to further cleavage of C3b by soluble inhibitors in serum (such as factor H and factor I) when the surface is coated with high densities of C3b (*27, 28*). We further tested the deposition of the C5b-9 complex. As a result of amplified complement cascades, significantly increased deposition of the complex was induced by SARS-CoV N protein at a much lower concentration (Fig. S2F), similar to that observed in patient sera (*22*).

Activated complement plays a crucial role in the efficient phagocytosis of pathogens and cellular debris by C3b or C5b-mediated opsonization (*29*). To study complement-dependent phagocytosis, *E. coli* and mouse peritoneal macrophages were incubated together in diluted serum with or without SARS-CoV N protein. Bound C3 or its large fragment C3b, the product after C3 cleavage, was stained with FITC-labeled anti-C3c antibody, and the FITC-positive macrophages containing C3-conjugated *E. coli* were counted under a microscope. In concert with complement activation, complement-dependent phagocytosis by mice peritoneal macrophage in the presence of mouse serum containing complement component including MASP-2 was enhanced remarkably by SARS-CoV N protein compared with the HSA control (Fig. S2G). These findings indicated that SARS-CoV N protein effectively promoted the activation and opsonic effect of the complement system.

### N proteins of SARS-CoV and MERS-CoV aggravates LPS-induced pneumonia by MASP-2-involved complement activation

Persistent activation of complement leads to uncontrolled inflammation. To investigate the effect of N protein-potentiated MASP-2 activation on inflammation, mice were pre-infected with adenovirus (1×10^9^ PFU) expressing SARS-CoV N (Ad-SARS N) or its mutant or adenovirus vehicle only (Ad-null). The mice were then challenged with LPS, which contains MBL-binding motifs, to activate LP and induce inflammation (*30*). 8 of 10 mice pre-exposed to adenovirus vehicle survived when challenged with 5 mg/kg LPS (Fig. 3A), while all 10 mice pre-exposed to Ad-SARS N died within 12 hr after LPS administration at the same dosage (Fig. 3A). Severe lung damage and massive inflammatory cell infiltration were also observed in dead mice (Fig. 3B). In concert with previous findings, the pre-infection of Ad-229E N and Ad-SARS N bearing the Δ116-124 or Δ321-323 deletion in the N protein, respectively, had significantly reduced effects on mouse mortality. Importantly, when anti-N, anti-MASP-2 antibody or C1INH was administered simultaneously with LPS administration in the Ad-SARS N pre-infected mice, both death rate and lung tissue inflammation induced by LPS were significantly reduced (Fig. 3A and 3B). These results collectively demonstrate that the SARS-CoV N protein greatly potentiated LPS-induced inflammation via MASP-2 activation, which thereby initiates LP-involved complement cascade activation.

**Fig 3.**
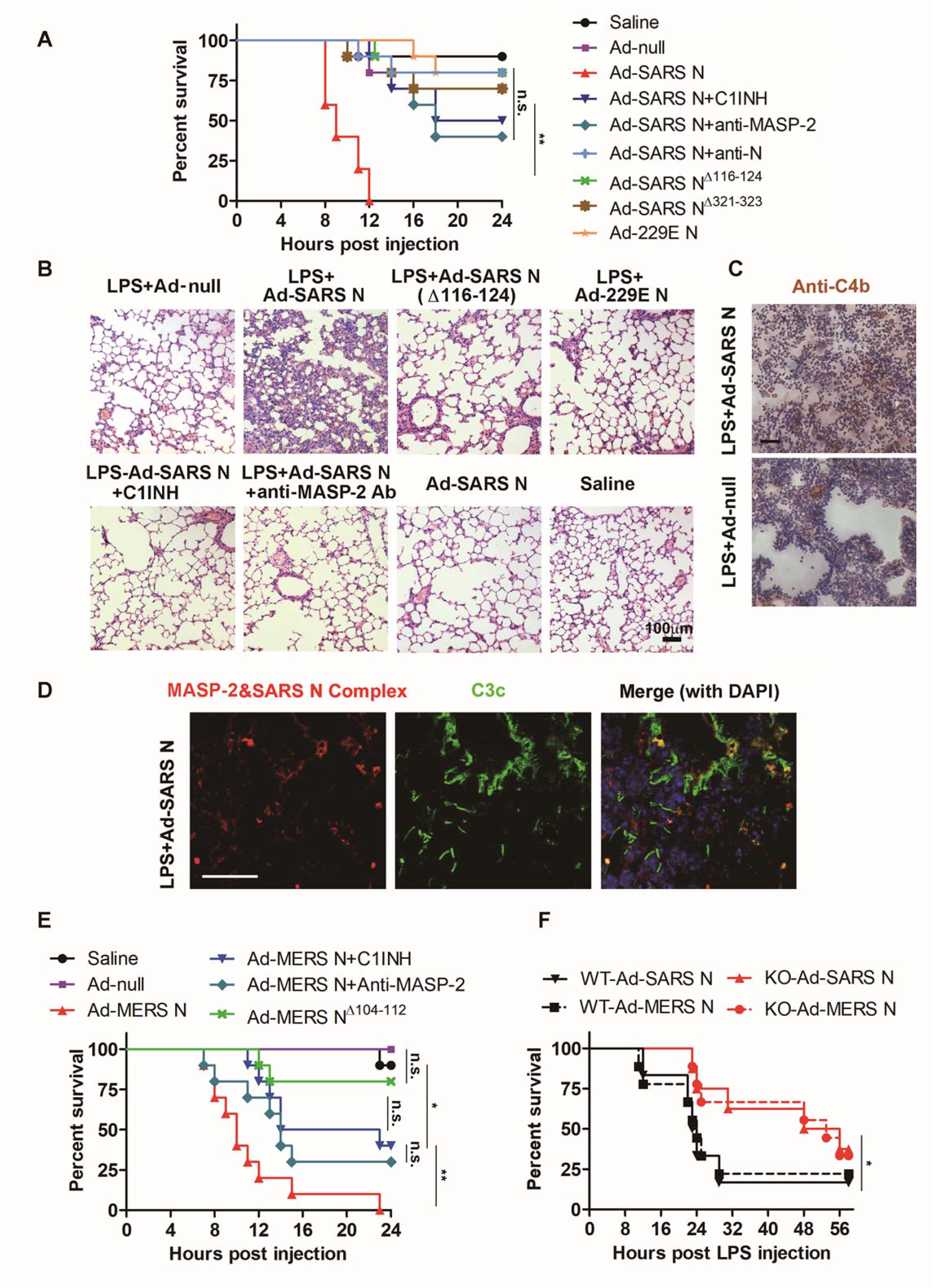
N proteins potentiate LPS-induced pneumonia in mice. (A) BALB/c mice (10 mice/group) were infected with 1×10^9^ PFU Ad-SARS N/Ad-null or a saline control, and LPS (5 mg/kg) was given on the 6th days post infection (d.p.i.). Antibodies (200 μg/kg) or C1INH (4 mg/kg) was injected 30 min before LPS injection. The fatality of mice was noted. (B) Lung paraffin sections were analyzed by HE staining. (C) Mice were infected with 1×10^8^ PFU Ad-SARS N/Ad-null, and LPS (5 mg/kg) was given by on 6 d.p.i. Mice were sacrificed after LPS challenged for 6 hr. Frozen lung sections were stained with anti-C4b antibody. (D) SARS-CoV N and MASP-2 complex formation in frozen lung sections was measured by *in situ* PLA, as indicated by the red signals. Deposited C3 fragments were stained with FITC-labeled anti-C3c antibodies (green). Scale bar, 50 μm. (E) Mice were pre-infected with 1×10^9^ PFU Ad-MERS N/Ad-null and treated with LPS, anti-MASP-2 antibody or C1INH as mentioned above. The mice survival curve was plotted. (F) *Masp2*^-/-^ and *Masp2*^+/+^ C57BL/6N mice were infected N-expressing adenovirus and injected with LPS as mentioned above. The mice survival curve was plotted.

To investigate the complement activation in lung of mice induced by LPS and N protein, mice pre-infected with Ad-SARS N (1×10^8^ PFU) were challenged with LPS (5 mg/kg). At 6 hr post LPS administration, the mice were sacrificed, and the paraformaldehyde-fixed lung tissue was subjected to immunohistochemical (IHC) staining with anti-C4b antibody or immunofluorescence analysis with FITC-labeled anti-C3c antibody. The C4b and activated C3 deposition in the lung were significantly increased in mice expressing SARS-CoV N protein (Fig. 3C and the middle panel of Fig. 3D), compared with the weak staining in the lungs of mice treated with LPS and Ad-null (Fig. 3C and the middle panel of Fig. S3A).

The *in vivo* association of MASP-2 with the N protein was further assessed by *in situ* proximity ligation assay (PLA) with anti-N and anti-MASP-2 antibodies (the left panels of Fig. 3D and S3A), and red signals, which were only detectable when SARS-CoV N and MASP-2 bind to each other, were abundantly observed in pulmonary cells from mice expressing SARS-

CoV N but not negative control animals (Fig. S3A). These results confirmed an *in situ* direct binding and activation of MASP-2 by SARS-CoV N in mice lung tissue.

Similarly, mice challenged with LPS suffered serious pneumonia and 100% die within 24 hr when pre-infected with adenovirus expressing MERS-CoV N protein (Ad-MERS N) but not its Δ104-112 mutant (Ad-MERS N^Δ104-112^) for 7 days, which was partially rescued by C1INH and anti-MASP-2 antibody (Fig. 3E). These results indicate that the N protein is employed by both SARS-CoV and MERS-CoV to promote complement activation through the MASP-2-mediated lectin pathway.

The pathogenicity of N protein was further investigated using *Masp2* knockout (KO) mice (KO region was indicated by Fig. S3B) with MASP-2 protease activity deficiency by the same way. Mice were pre-infected with Ad-SARS N or Ad-MERS N (1×10^9^ PFU) for 5 days, and then challenged with LPS. Compared with wild-type mice, *Masp2* KO mice survived longer and had a higher survival rate (Fig. 3F), which may be attributed to a compromised complement activation because of MASP-2 deficiency.

### Complements cascade is over-activated in lungs of COVID-19 patients

Because the SARS-CoV and SARS-CoV-2 induce only mild lung damage in mice, and investigation with live SARS-CoV, including mouse adapted SARS-CoV, is not allowed by related regulations in China, we try to get the clinical evidence that the over-activation of complement LP pathway occurred in COVID-19 patients. The paraformaldehyde-fixed lung tissue of patients who died of COVID-19 were collected and subjected to IHC staining with MBL, MASP-2, C4alpha, C3 or C5b-9 antibodies. All these complement cascade components demonstrated strong IHC staining signals in the patient lung tissue (Fig. 4A-4E), which suggested that the complement components were deposited in type I and type II alveolar epithelia cells, as well as inflammatory cells, some hyperplastic pneumocytes, and exudates in alveolar spaces with necrotic cell debris. Notably, SARS-CoV-2 N protein was also observed in the lung tissue of died patients by autopsy (*31*). Further, significantly increased serum C5a level was also observed in COVID-19 patient, particularly in severe cases (Fig. 4F). These results collectively indicated that complement pathways were aggressively activated in the lungs of COVID-19 patients, which may be attributed to SARS-CoV-2 N protein.

**Fig 4.**
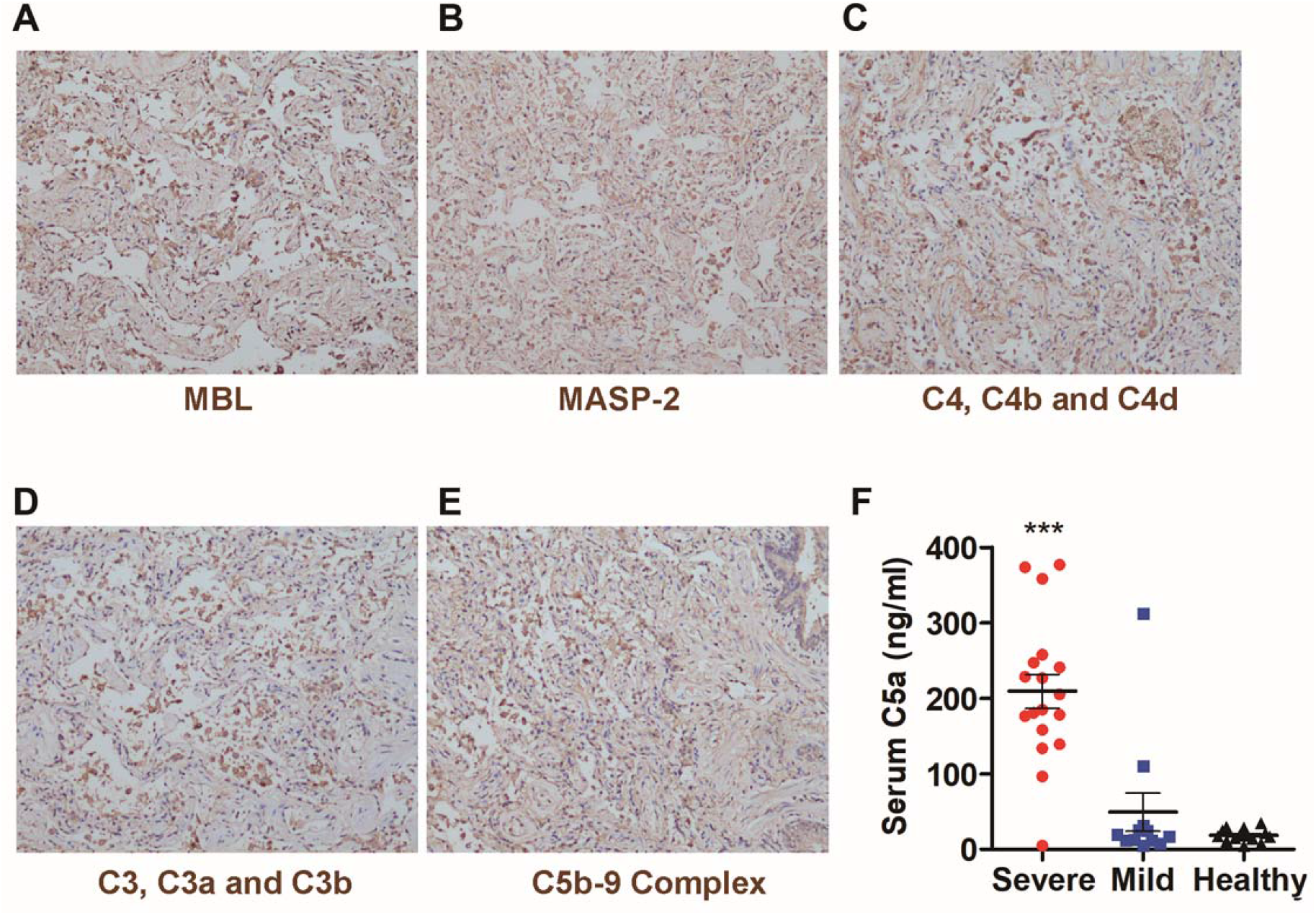
Complements activation in COVID-19 patients. (A-E) Paraformaldehyde-fixed lung tissues from postmortem autopsy was used for paraffin tissue sections and IHC staining with indicated antibodies. Microphotography was carried out by Olympus BX52 microscope under a 10× objective. (F) Serum C5a from healthy people, mild or severe COVID-19 patients were analyzed by ELISA.

### Complement-targeted therapy shows a promising curative effect against COVID-19

It has been widely accepted that the excessive inflammatory and cytokine storm greatly contribute to the severity and lethality of COVID-19, which may be attributed to the unrestrained activation of complement pathway. Therefore, downregulation of MASP-2 as well as its downstream signal molecules, such as the potent anaphylatoxin C5a, may provide a new approach to control the pneumonia induced by the SARS-CoV-2.

Based on our finding and its potential and tempting application in COVID-19 therapy, a recombinant C5a antibody (BDB-001 injection by Staidson (Beijing) Biopharmaceuticals Co., Ltd), which was in phase I clinical trial for hidradenitis suppurativa, was rapidly approved by National Medical Product Administration (NMPA) for clinical trials for the treatment of COVID-19 (2020L00003). “A multicenter, randomized double blind placebo-controlled trial in mild COVID-19 patients” and an open label “two-cohort clinical trial in patients with severe and critical COVID-19” were carried out simultaneously under the approval from ethic committee of Huoshensan Hospital with the informed consent of the patients. While the larger dataset from more patients in the COVID-19 cohort will be reported elsewhere when the trial is finished, here we reported the first 2 cases administrated with anti-C5a antibody in the open label trial.

Patient #1, a 54 years old male resident in the city of Wuhan, was admitted to Dongxihu Hospital on the 4th day after the onset of symptoms (5 days of illness) with fever. Infection of SARS-CoV-2 was confirmed by rRT-PCR for SARS-CoV-2, and chest CT scan showed bilateral opacities. The disease was getting worse from day 9 of illness, with high fever (38.7°C∼39.8°C), the percutaneous oxygen saturation (SpO2) <93% on room air, pneumonia progression on CT scan, and severe hepatic damage. Prednisone (40 mg/day for 4 days) was administrated on day 10 through 13 of illness but the condition deteriorated, so the patient was transferred to Wuhan Huoshenshan Hospital, a new hospital established urgently for severe COVID-19 patients, on the day 14 of illness (Fig. 5A), and was considered as severe case in critical condition with moderate ARDS (PaO2/FiO2 ratio <150), respiratory rate 30/min,and SpO2 dropped to 77% when exposed to room air for 1 minute. Supportive care was provided, including high-flow nasal oxygen (HFNO) to target SpO2>95% (Fig. 5B, left). He was also administrated with the antibiotics (moxifloxacin) and human serum albumin, while prednisone was discontinued. Treatment with anti-C5a monoclonal antibody (BDB-001) was initiated on the morning of day 15 of illness. The antibody was given intravenously in 250 ml saline, at a dosage of 300 mg/day, on day 1, 2, 3, 5, 7, 9, 11 and 13. No adverse event was observed and the clinical condition improved in the next days, with normal body temperature in the evening of the same day (Fig. 5C, left), increased PaO2/FiO2 ratio (Fig. 5B, left) and lymphocyte cell number (Fig. 5C, left), decreased C reactive protein (CRP) concentration (Fig. 5C, left), and significantly improved hepatic function (shown by decreased ALT, AST, and increased total serum protein and serum albumin concentration) (Fig. 5D, left). The fraction of inspiration O_2_ and the gas flow rate of HFNO was eventually decreased from the highest (80%, 40 L/min to 30%, 20 L/min) to target a SpO2>95% (Fig. 5B, left). Because the high risk of taking the patient in critical condition to CT scan in the temporarily established hospital (the long distance and the cold raining weather without HFNO), no CT scan just before anti-C5a administration was available for evaluation. Nevertheless, the pneumonia 20 days after the 1st dose is observed obviously improved than 11 days after the 1st dose (Fig. S4, upper panel).

**Fig 5.**
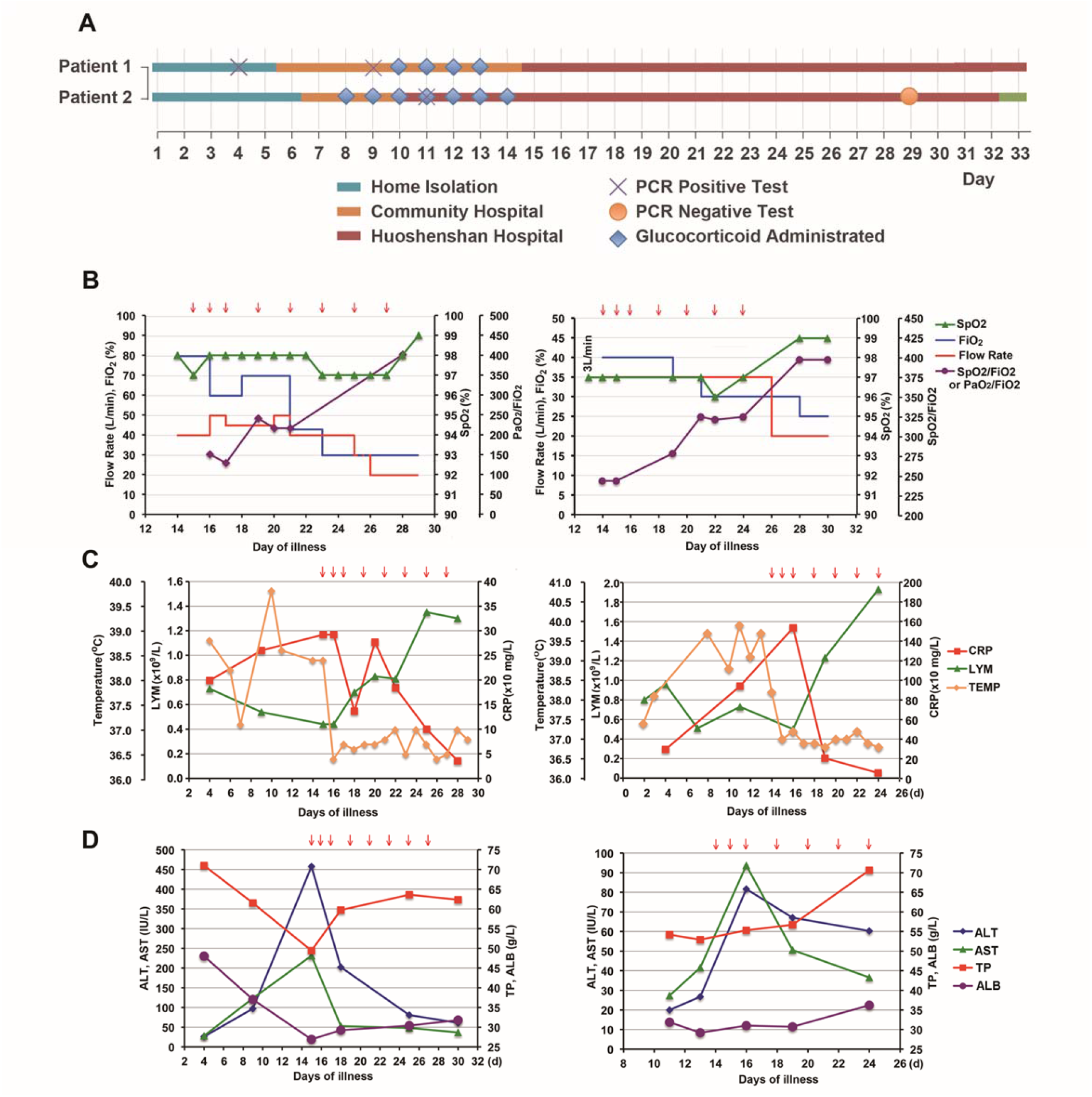
Treatment of COVID-19 patients with anti-C5a antibody. (A) Timeline of illness onset, SARS-CoV-2 RNA detection and hospitalization of the two patients. (B) Flow rate, fraction of inspiration (FiO_2_) of high-flow nasal oxygen, percutaneous oxygen saturation (SpO2) and PaO2/FiO2 or SpO2/FiO2 ratio in patient #1 (Left) and patient #2 (Right). (C) Body temperature (TEMP), C reactive protein (CRP) level and blood lymphocyte number (LYM) changes in patient #1 (Left) and patient #2 (Right). (D) Hepatic function changes in patient #1 (Left) and patient #2 (Right). ALT: alanine aminotransferase; AST: aspartate aminotransferase; TP: total protein; ALB: albumin.

Patient #2 was a 67 years old male who was admitted to the Sixth Hospital of Wuhan on the 5th day after the onset of symptoms (6 day of illness) with fever and cough. CT scan showed opacity on the superior lobe of left lung (Fig. S4). Infection of SARS-CoV-2 was also confirmed by rRT-PCR at day 11 of illness. Anti-viral (Arbidol) and antibiotics (moxifloxacin) was administrated together with other supportive treatment. Condition was worsening by day 8 with severe cough and high fever (39.7°C) (Fig. 5C, right). Methylprednisolone (40 mg/day, for 7 days from the 8th day after illness) showed little if any improvement of symptoms. The patient was transferred to Wuhan Huoshenshan Hospital on day 10 (Fig. 5A) and the illness continued getting worse as shown by low SpO2 (<90% on room air on day 14), high fever (>39°C on day 10-13) (Fig. 5C, right) and pneumonia on chest CT on day 11 of illness (2 days before the 1st dose) (Fig. S4, lower panel). The patients reported severe cough, chest tightness and dyspnea. HFNO has to be given to maintain SpO2>95%. Anti-C5a was administrated on the morning of day 14 of illness, and continued as in the patient #1. Subsequently, a normal temperature was observed on the same day (Fig. 5C, right). Cough, dyspnea and oppression in chest were reported to be getting better the next day (Day 15 of illness). A significant decreased CRP level, significantly increased white blood cell and lymphocyte number were also observed in the next days (Fig. 5C, right). Hepatic function was gradually improved (Fig. 5D, right, data on day 15 are not available). Flow rate and Fraction of inspiration O2 necessary to maintain SpO2>95% gradually decreased, with the increasing SpO2/FiO2 ratio (Fig. 5B, right). Chest CT on day 26 of illness (12 days after the 1st dose) also showed alleviated pneumonia (Fig. S4, lower panel).

These data provided the first evidence for a damaging role of C5a in severely affected COVID-19 patients. The two severe patients are significantly benefited from anti-C5a monoclonal antibody therapy, which suggested that an aberrant complement cascade should be considered as a promising therapeutic target of COVID-19.

## Discussion

In the past 17 years, successively emerged SARS-CoV, MERS-CoV and SARS-CoV-2 broke through the species barrier and brought new infectious diseases and social panic to human. All these highly pathogenic coronavirus cause acute lung injury and acute respiratory distress syndrome (ARDS), therefore resulting in an increased severity and lethality. Patients infected with the virus may develop to atypical pneumonia resulting from severe immune injury. The excessive human immune responses that were characterized by the extensive release of pro-inflammatory cytokines and chemokines, called a “cytokine storm”(*5*), is thought to be the major initiator of the sever pneumonia caused by high pathogenic coronavirus, including the latest SARS-CoV-2 (*4*). Although immunopathogenesis has been partially implicated in SARS or MERS, the mechanism responsible for virus-induced hyper-activation of host immune system remains poorly understood.

Here we show that SARS-CoV, MERS-CoV and SARS-CoV-2 share a common mechanism connecting the viral N proteins to binding and potentiation of an MBL, Ca^2+^-dependent auto-activation of MASP-2, leading to the uncontrolled activation of complement cascade, as characterized by the enhanced C4 cleavage and complement deposition (Fig. 1 and 2). The binding of N protein to MASP-2 amplifies the effects of MASP-2-mediated lectin pathway activation. N protein mutants either failed to interact with MASP-2 or failed to form an N dimer have little or no effect on MASP-2-mediated lectin pathway activation (Fig. 1C and 1D), suggesting that the effects of N protein are dependent on binding and dimerization.

As a “dual-edged sword”, complement is critical for innate immunity against pathogens. Anaphylatoxins, such as C3a and C5a, can activate immune cells, and therefore induce the release of various cytokines. Activated complement cascade produces the cytolytic terminal complement complex C5b-9 and the C3b and C5b fragments (*29, 32*). These peptides induce the synthesis of arachidonic acid metabolites, including prostaglandin (PG) E2, thromboxane B2 and leukotrienes (*30, 33*), which further induce the recruitment and activation of neutrophils, monocytes and eosinophils, and stimulate the production of a number of pro-inflammatory cytokines and mediators (*34*). These cytokines trigger and maintain inflammatory processes and help the innate immunity to fight the virus. Nevertheless, complement-involved innate immunity activation must be fine-tuned because unrestrained complement activation always contributes to disseminated intravascular coagulation (DIC), inflammation, cell death, and immune paralysis and ultimately leads to multiple organ failure and death. The finding that the coronavirus N protein potentiating complement activation provides a new insight into the causation of pneumonia induced by SARS-CoV, MERS-CoV and SARS-CoV-2 infection. Additionally, we also noticed that the pre-infection of Ad-SARS N or Ad-MERS N evidently increased the fatality of LPS-induced pneumonia (Fig. 3), underlining the potential role of N protein in the inflammatory injury of tissue caused by the massive LPS released from secondary bacterial infections (*35, 36*).

The involvement of N protein-mediated MASP-2 and therefore the complement cascade over-activation in the pathogenesis of coronavirus provides new potential strategies to develop therapeutics against SARS, MERS or the latest COVID-19, a schematic summary of which was demonstrated in Fig. 6.

**Fig 6.**
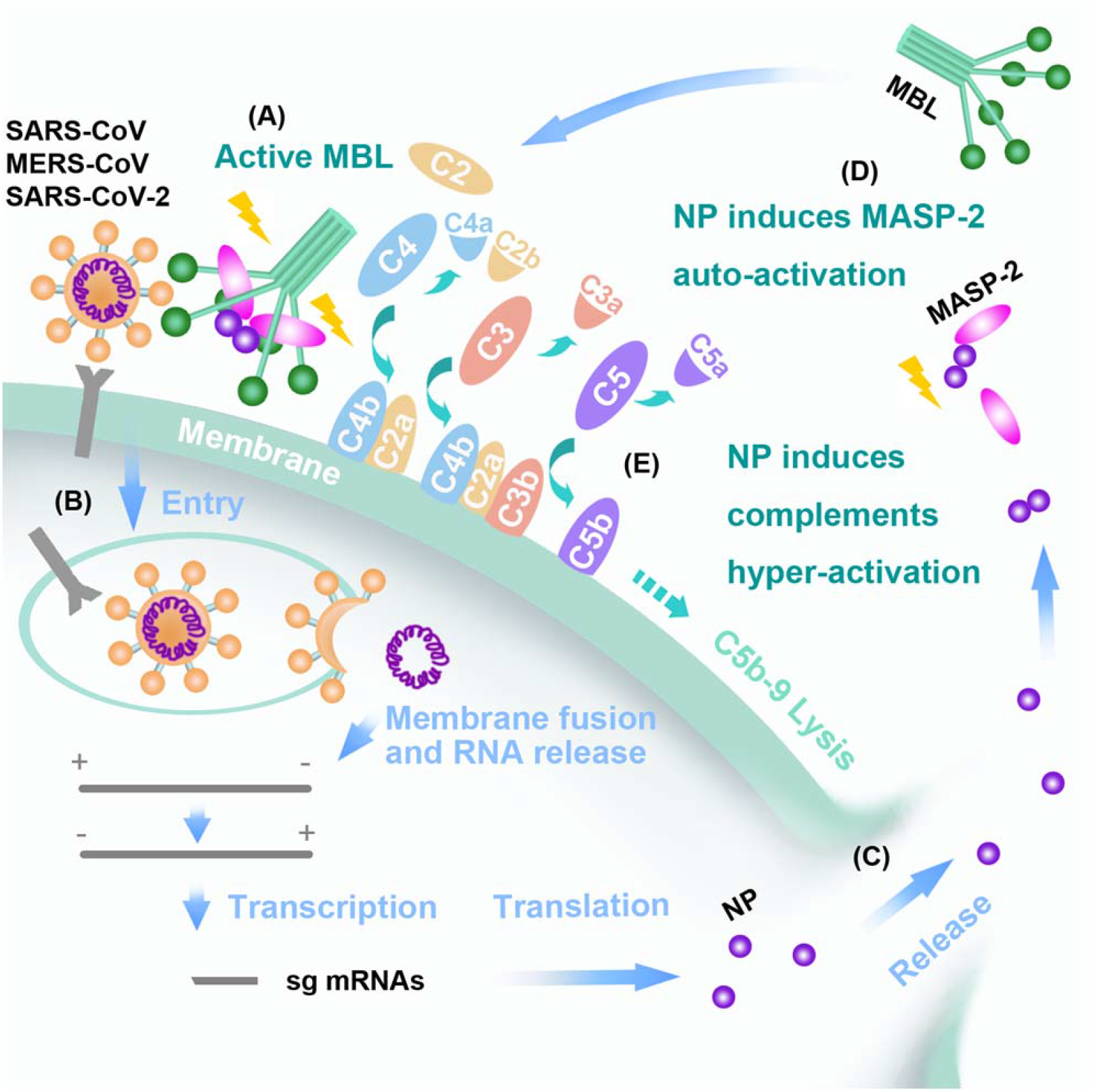
Schematic representation of MBL pathway over-activated by N protein of SARS/ MERS-CoV or SARS-CoV-2. (A) Virus binds to cell surface and S protein actives MBL. (B) Virus enters cells and expresses viral proteins including N protein. (C) N proteins release by secretion or after cells and virus lysis. (D) The extracellular soluble N protein dimmers interact with MASP-2, induce MASP-2 auto-activation and binding to MBL. (E) The accelerated activation of MASP-2 induces downstream complements cascades over-activation of the MBL pathway, and promotes cell lysis and N protein releases by secretion or complement mediated cytotoxicity, which may result in uncontrolled tissue damage and inflammation.

Firstly, neutralization of N protein in the serum by antibody effectively alleviated lung injury and reduced the fatality in LPS/Ad-SARS N challenged mice. Coincidentally, SARS patients who produce higher levels of N protein specific but not spike protein specific antibodies tend to recover more rapidly, suggesting that levels of N antibodies correlated with outcome of SARS (*37*). According to our findings, similar mechanisms may be present in MERS-CoV or SARS-CoV-2 infection.

Secondly, *Masp2* KO mice showed significantly mild symptoms and a shorter course of disease in LPS induced, N protein boosted mice pneumonia model, which confirmed the harmful role of MASP-2 in this severe pneumonia. Accordingly, administration of anti-MASP-2 antibody or the MASP-2 inhibitor C1INH showed a promising treatment effect (Fig. 3). Improved MASP-2 antibodies that have higher affinity and neutralization activity, such as OMS721 (*38*) (which showed promising effects on thrombotic microangiopathy), or an injectable C1INH medicine, such as HAEGARDA, may provide effective protection. Further clinical evaluation is of value to be carried out for the treatment of SARS-CoV-2 induced pneumonia.

Thirdly, we’ve observed an excessive activation of complement cascade in lung tissue of dead patient, which is coincident with the observation in Ad-SARS N pre-infected and LPS-primed mice model. High-level C5a was accumulated in the serum of severe but not mild COVID-19 patients (Fig. 3 and 4). So, it will make sense that complement cascade-targeted immunomodulation may be effective for inflammation-control in highly pathogenic coronavirus-related diseases. In this pathway, C5a is the most potent complement protein triggering inflammation. Blockage of C5a signaling has also been reported to improve treatment of acute lung injury (ALI) induced by highly pathogenic viruses (*39*). Based on our scientific finding and the good safety record in phase II trial for other disease by Staidson and InflaRx GmbH (whom produce the anti-C5a antibody in the same cell line), the recombinant anti-C5a antibody produced by Staidson Biopharmaceuticals was approved by NMPA for the clinical trial for treatment of severe and critical COVID-19 patients. At least in the first two patients, both are in the disease deterioration, the anti-C5a antibody showed rapid and promising effect exceeding the expect of clinical physicians. Although the final efficacy will be released until the clinical trial is finished, it is worth to expect that anti-C5a antibody would provide a new approach for the treatment of COVID-19.

## Materials and Methods

### Cell culture and transfections

The 293T cell line was obtained from the Cell Resource Center of Peking Union Medical College. Cells were grown in Dulbecco’s modified Eagle’s medium (DMEM) (Invitrogen) supplemented with 10% heat-inactivated fetal bovine serum (FBS) (HyClone), 2 mM L-glutamine, 100 units/ml penicillin, and 100 µg/ml streptomycin. Cells were transfected with plasmid DNA using Lipofectamine 2000 (Invitrogen) according to the manufacturer’s protocol.

### Vectors and epitope tagging of proteins

The N gene of SARS-CoV (GenBank Accession #AY274119) was amplified by RT-PCR from the SARS-CoV RNA of patient serum samples (Upstream primer: 5’-CGGAATTCCATATGTCTGATAATGGACCCCAA-3’; downstream primer: 5’-CGGGATCCTTATGCCTGAGTTGAATCAGC-3’) and cloned into the pcDNA3-based Flag vector (Invitrogen), pCMV-Myc (Clontech), pGEX-4T-2 (GE Health care), and the BglII and EcoRI sites of pEGFPC1 (Clontech). The N gene of MERS-CoV was chemically synthesized (Huaxinrcomm Technology Co., Ltd) and cloned into the pcDNA3-based Flag vector at the BamHI and EcoRI sites. The N gene of SARS-CoV-2 was chemically synthesized (General Biosystems (Anhui) Co. Ltd) and cloned into the pcDNA3.1-based HA vector at the KpnI and XbaI sites. The *MASP2* gene was amplified from human hepatocyte cDNA library and inserted into corresponding vectors.

### Immunoprecipitation and immunoblotting

Cell lysates were prepared in lysis buffer (50 mM Tris-HCl, pH 7.5, 1 mM phenylmethylsulfonyl fluoride, 1 mM dithiothreitol, 10 mM sodium fluoride, 10 µg/ml aprotinin, 10 µg/ml leupeptin, and 10 µg/ml pepstatin A) containing 1% Nonidet P-40. Soluble proteins were subjected to immunoprecipitation with anti-Flag M2 agarose (Sigma). The adsorbates were then separated by SDS-PAGE and transferred onto an Immobilon-P transfer membrane (Millipore) by semi-dry transblot (Biorad). The membrane was blocked by 5% Western-Blocker (Biorad). Immunoblot analysis was performed with horseradish peroxidase (HRP)-conjugated anti-Flag (Sigma), anti-β-actin (Sigma), anti-green fluorescent protein (GFP) (Clontech), anti-MASP-2 (Santa Cruz), anti-C4α (Santa Cruz), HRP-conjugated anti-Myc (Santa Cruz), and goat anti-mouse immunoglobulin G (IgG) (Amersham/Pharmacia) antibodies. The antigen-antibody complexes were visualized by chemiluminescence (GE Health Care).

### Purification of SARS-CoV and MERS-CoV N protein

As described previously (*40*), pET22b-SARS/MERS-CoV N was transformed into the expression strain BL21 (DE3). After induction with 1 mM IPTG for 8 h, the bacteria were harvested by centrifugation and re-suspended in buffer A (25 mM Na_2_HPO_4_/NaH_2_PO_4_ (pH 8.0), 1 mM EDTA, and 1 mM DTT) before sonication. Soluble N protein in the lysate was purified with ion-exchange chromatography with SP-Sepharose Fast Flow (25 mM Na_2_HPO_4_/NaH_2_PO_4_ (pH 8.0), 1 mM EDTA, 1 mM DTT, and 0.35-0.5 M NaCl), followed by Superdex 200 gel filtration (GE Healthcare) and elution with buffer A. *E. coli* transformed with the vector pET22b was lysed as described above, and the eluate was used as a negative control for the purified N protein.Purified SARS-CoV-2 N-His was obtained from General Biosystems (Anhui) Co. Ltd.

### Purification and renaturation of MASP-2

Recombinant protein expression and renaturation were performed as described (*24, 41*). In brief, pET22b-MASP-2 was transformed into the expression host strain BL21 (DE3). After induction with 1 mM IPTG, cells were harvested and sonicated. The inclusion bodies were solubilized in 6 M GuHCl, 0.1 M Tris-HCl (pH 8.3), and 100 mM DTT at room temperature; the solubilized proteins were then diluted into refolding buffers containing 50 mM Tris-HCl, 3 mM reduced glutathione (Sigma), 1 mM oxidized glutathione (Sigma), 5 mM EDTA, and 0.5 M arginine. The protein samples were then cooled to 4°C. The renatured protein was dialyzed against 20 mM Tris, 140 mM NaCl, pH 7.4 at 4°C, concentrated with PEG8000, aliquoted, and stored at −70°C.

To obtain high-activity MASP-2, Flag-tagged MASP-2 was expressed in 293T cells, precipitated with anti-FLAG magnetic beads, and eluted with Flag peptide (Sigma). The concentration of MASP-2 was assessed using a BCA kit and immunoblot analysis, with the purified prokaryotic-expressed MASP-2 as a standard control.

### MASP-2 auto-activation and C4 cleavage assay

Purified MASP-2 (8 nM) was incubated at 37°C in 20 mM Tris-HCl (pH 7.4), 150 mM NaCl, and 2 mM CaCl_2_ with purified C4 (Calbiochem), recombined MBL (Calbiochem), mannan (Sigma), and SARS-CoV N protein at concentrations of 50 nM, 30 nM, 15 ng/ml, and 10 nM, respectively. The cleavage was followed by SDS-PAGE under reducing conditions, and the C4alpha’ fragments, which are left by C4a release after C4 cleavage and can indicate C4 activation, and MASP-2 were detected by immunoblot analysis with anti-C4alpha chain antibody (Santa Cruz) or anti-Flag antibody (Sigma). C4b containing C4alpha’ but not C4a was used to indicate C4alpha’ band position.

### Complement deposition assay

The C4b deposition assay was performed using a human MBL/MASP-2 assay kit (Hycult biotech) (*42*). In brief, diluted serum was incubated in mannan-coated plates with high salt binding buffer overnight at 4°C and removed by washing, and the MBL-MASP-2 complex was captured. Purified C4 and N protein were added and incubated for 1.5 hr, and the deposited C4b was detected following standard protocols. The functional activity of LP and AP was assessed by ELISA as previously described (*43*). Nunc Maxisorb plates were coated with 10 μg mannan per well in 100 mM Na_2_CO_3_/NaHCO_3_ (pH 9.6) at room temperature overnight. After each step, plates were washed three times with PBST (300 μl/well). Residual binding sites were blocked by incubation with 10 mM Tris-HCl (pH 7.4), 150 mM NaCl, and 2% HSA for 2-3 hr at room temperature. Serum samples were diluted 1:80 in 10 mM Tris-HCl (pH 7.4) containing 150 mM NaCl, 0.5 mM MgCl_2_, 0.05% Tween-20, and 0.1% gelatin with or without 2 mM CaCl_2_ and N protein. All samples and buffers were prepared on ice. The plates were then sequentially incubated for 1 hr at 4°C and for 1.5 hr at 37°C followed by washing. All incubation volumes were 100 μl. Complement binding was detected using antibodies followed by washing. Detection of C4, activated C3, and C5b-9 was performed using anti-C4α chain antibody (Santa Cruz), anti-activated C3 antibody (Santa Cruz), and anti-C5b-9 antibody (Calbiochem), respectively. Antibody binding was detected using HRP-conjugated sheep anti-mouse antibody or donkey anti-rabbit antibody (R&D). Enzyme activity of HRP was detected using TMB incubation for 30-60 min RT, and the reaction was stopped with 2 M H_2_SO_4_. The OD was measured at 450 nm using a microplate reader.

### MASP-2:MBL binding assay

Binding of MASP-2 to MBL was assessed by ELISA. As mentioned above, Nunc Maxisorb plates were coated with 10 μg mannan per well in 100 mM Na_2_CO_3_/NaHCO_3_ (pH 9.6) at room temperature overnight and blocked with 2% HSA. MBL protein (1 μg/ml) was incubated in 10 mM Tris-HCl (pH 7.4), 150 mM NaCl, 5 mM CaCl_2_, 100 μg/ml HSA, and 0.5‰ TritonX-100 at 4°C for 2 hr. Purified MASP-2 and N (or control) proteins were added to the wells at different times to obtain final concentrations of 0.2 mg/ml and 200 ng/ml, respectively. The plates were washed after 32 hr of incubation at 4°C, and the binding of MASP-2 were detected with anti-MASP-2 antibody followed by HRP-conjugated rabbit anti-goat antibody. The enzyme activity of HRP was detected using TMB incubation for 30-60 min at RT, and the reaction was stopped with 2 M H_2_SO_4_. The OD was measured at 450 nm using a microplate reader.

### Opsonocytophagic assay

Mouse cells isolated from peritoneal cavity were washed and inoculated with RPMI 1640 Media (10% FBS) in 96-well plates for 2 hr at 37°C. Serum was diluted by 0.781%, 1.562%, 3.125%, 6.25%, 12.5%, 25%, 50% and 100% with 1×PBS, 1mM CaCl_2_, and 2mM MgCl_2_. Diluted serum, SARS-CoV N protein (100ng/ml) and *E. coli* (the ratio to cells was 10:1) were added to each well and incubated for 30 minutes at 37°C. The eluate from pET22b-transformed *E. coli* was used as a negative control for purified N protein to exclude effects due to bacterial components that may active complement. The reaction was stop and cells were fixed with 10% neutral formalin. Complement C3c depositon was detected with FITC-C3c antibody, and the stained cells were counted. The points represent the mean values from two repeated wells. Error bars, mean±S.D. *P<0.05 and **P<0.01 by unpaired two-tailed Student’s t-test.

### Animal experiments

Groups of BALB/c mice were provided from the experimental animal center of the Academy of Military Medical Sciences. *Masp2*^-/-^ (KO) mice and littermate control wild type (WT) mice were provided from Cyagen Biosciences Inc. All mice were maintained in the experimental animal center of the Academy of Military Medical Sciences (China). Mice (8-10/group) were infected three times (day 1, 2, 3) with 1×10^8-9^ PFU Ad-N/Ad-null (Beijing BAC Biological Technologies) or a saline control via the tail vein, and LPS (5 mg/kg) was given via the tail vein on the 5-6th day. Anti-MASP-2 monoclonal antibody (200 μg/kg, HBT), anti-N monoclonal antibody (200 μg/kg, Sino Biolgical) or C1INH (4 mg/kg, Calbiochem) was injected via the tail vein 30 min before LPS injection. This study was performed with the approval of the ethics committee at the Beijing Institute of Biotechnology and conformed to the relevant regulatory standards.

### Immunohistochemistry

Postmortem autopsy from the 4 patients died in Huoshenshan hospital was carried out by Dr. Xiuwu Bian under the approval of the hospital ethic committee and the family member of the death. The detailed results will be published elsewhere. Paraformaldehyde-fixed lung tissues was used for paraffin tissue sections and immunohistochemical staining with MBL (Santa Cruz), MASP-2 (Santa Cruz), C4α chain (Santa Cruz), C3 (Santa Cruz) or C5b-9 (Calbiochem) antibodies as described in the instruction manual.

### Detection of C5a in COVID-19 patients

Sera from mild or severe COVID-19 patients were collected under the approval of hospital ethic committee. Server patients were defined as fever or suspected respiratory infection, plus one of respiratory rate >30 breaths/min, severe respiratory distress, or SpO2 <90% on room air. Patients with pneumonia and no signs of severe pneumonia are defined as mild cases. Sera from 12 mild patients (6 males and 6 females), with an average of 56±12.1 years old and an average 26.3±9.1 days of illness, and sera from 18 severe patients (7 males and 11 females), with an average of 72.4±10.1 years old and an average 40.15±12.74 days of illness, were assayed. Sera from healthy people for physical were collected from the clinical laboratory. Serum C5a level was detected by double antibody sandwich ELISA (R&D Systems).

### Anti-C5a antibody therapy

BDB-001 is a mouse-human chimeric (IgG4) antibody against human C5a. It was manufactured from IFX-1 cell line by Staidson in accordance with the Good Manufacture Practices. The IFX-1 cell line has originally been developed by InflaRx GmbH, Germany and was licensed to Beijing Deferengrei Biotech Co., Ltd., a wholly owned subsidiary of Staidson Biopharmaceutical Co., Ltd. The injection was approved by National Medical Product Administration (NMPA) of China for phase I clinical trial in healthy subjects in 2018. After the SARS-CoV-2 coronavirus induced pneumonia (COVID-19) outbreak, the antibody was approved by NMPA for phase II clinical trials for the treatment of COVID-19 on February 7, 2020 (2020L00003). Administration of the antibody to human was also approved by the Ethics Committee in Wuhan Huoshenshan Hospital. 300 mg anti-C5a antibody in 250 ml saline was administrated i.v. on day 1, 2, 3, 5, 7, 9, 11 and 13. Arterial blood gas (ABG) test, C reaction protein (CRP), blood routine (blood RT) and Hepatic function were regularly determined. SaO2, blood pressure, heart beat were monitored as required.

### Statistics

The data were presented as the mean ±SD or the mean ±SEM and analyzed using the two-tailed Student t test or ANOVA. Survival curves were analyzed by Log-rank test, Gehan-Breslow-Wilcoxon test or parametric regression model. Differences were considered significant at *P <0.05, **P <0.01, or ***P<0.001 as indicated.

## Data Availability

All data is always available for researches interested in this study.

## Acknowledgments

The authors wish to thank Dr. David Weaver for the help in the preparation of the manuscript.

## Author contributions

Conceived and designed the experiments: TG, XL, QM, XB, CC; Performed the experiments: TG, LZ, HL, QD, YJ, YF, XL, JL; Analyzed the data: TG CC YH; Contributed reagents /materials /analysis tools: WY, YL, HZ, PL, YL, QM, WC; Analyzed Clinical samples: XZ, ZZ, ZW, KL, HL; Performed clinical therapy: QM, SZ, YX, SL, LL, JP, LZ, LZ, MH, XC, YX, JS; Wrote the paper: TG, CC, XL;

## Data and materials availability

All data is available in the main text or the supplementary materials.

## Supplementary Materials

Figures S1-S4

## References

1. World Health Organization Multicentre Collaborative Network for Severe Acute Respiratory Syndrome Diagnosis, Lancet 361, 1730 (May 17, 2003).

2. N. Lee et al., N. Engl. J. Med. 348, 1986 (2003).

3. J. F. Chan et al., Clin Microbiol Rev 28, 465 (Apr, 2015).

4. C. Huang et al., The Lancet, (2020).

5. R. L. Graham, E. F. Donaldson, R. S. Baric, Nat. Rev. Microbiol. 11, 836 (Dec, 2013).

6. P. A. Rota et al., Science 300, 1394 (May 30, 2003).

7. J. F. Chan et al., Clin. Microbiol. Rev. 28, 465 (Apr, 2015).

8. P. Zhou et al., Nature, (2020).

9. M. Surjit et al., Journal of virology 79, 11476 (Sep, 2005).

10. F. Yasui et al., J. Immunol. 181, 6337 (Nov 1, 2008).

11. D. Deming et al., PLoS Med 3, e525 (Dec, 2006).

12. S. Sun et al., Am. J. Respir. Cell Mol. Biol. 49, 221 (Aug, 2013).

13. J. Zhou et al., J. Infect. Dis. 209, 1331 (May 1, 2014).

14. M. Wills-Karp, Proc. Am. Thorac. Soc. 4, 247 (Jul, 2007).

15. K. Takahashi, W. E. Ip, I. C. Michelow, R. A. Ezekowitz, Curr. Opin. Immunol. 18, 16 (Feb, 2006).

16. R. Wallis, Immunobiology 212, 289 (2007).

17. W. K. Ip et al., J. Infect. Dis. 191, 1697 (May 15, 2005).

18. Y. Zhou et al., Journal of virology 84, 8753 (Sep, 2010).

19. J. H. Chen et al., Proc. Natl. Acad. Sci. USA 101, 17039 (2004).

20. R. T. Pang et al., Clin. Chem. 52, 421 (Mar, 2006).

21. J. L. Liu, C. Cao, Q. J. Ma, Xi Bao Yu Fen Zi Mian Yi Xue Za Zhi 25, 777 (Sep, 2009).

22. X. Y. Che et al., Emerg Infect Dis 10, 1947 (Nov, 2004).

23. P. Gal et al., J Biol Chem 280, 33435 (Sep 30, 2005).

24. T. Gao, H. Zhao, X. Liu, C. Cao, Letters in Biotechnology 22, 806 (2011).

25. L. Beinrohr, J. Dobo, P. Zavodszky, P. Gal, Trends Mol. Med. 14, 511 (Dec, 2008).

26. S. V. Petersen, S. Thiel, L. Jensen, R. Steffensen, J. C. Jensenius, J Immunol Methods 257, 107 (Nov 1, 2001).

27. N. Rawal, R. Rajagopalan, V. P. Salvi, J. Biol. Chem. 283, 7853 (Mar 21, 2008).

28. M. Okroj, E. Holmquist, B. C. King, A. M. Blom, PLoS One 7, e47245 (2012).

29. D. S. Cole, B. P. Morgan, Clin. Sci. (Lond) 104, 455 (May, 2003).

30. M. Devyatyarova-Johnson et al., Infect. Immun. 68, 3894 (Jul, 2000).

31. X. Yao et al., Chin J Pathol 49, Epub ahead of print (2020).

32. J. R. Dunkelberger, W. C. Song, Cell Res. 20, 34 (Jan, 2010).

33. D. K. Imagawa, N. E. Osifchin, W. A. Paznekas, M. L. Shin, M. M. Mayer, Proc. Natl. Acad. Sci. USA 80, 6647 (Nov, 1983).

34. E. Sanchez-Galan et al., Cardiovasc. Res. 81, 216 (Jan 1, 2009).

35. C. Drosten et al., N. Engl. J. Med. 348, 1967 (May 15, 2003).

36. Y. Ami et al., Microbiol. Immunol. 52, 118 (Feb, 2008).

37. L. Zhang et al., J Med Virol 78, 1 (Jan, 2006).

38. W. J. Schwaeble et al., Proc. Natl. Acad. Sci. USA 108, 7523 (May 3, 2011).

39. R. Wang, H. Xiao, R. Guo, Y. Li, B. Shen, Emerg. Microbes Infect. 4, e28 (May, 2015).

40. B. Zhou et al., Journal of virology 82, 6962 (Jul, 2008).

41. G. Ambrus et al., J. Immunol. 170, 1374 (Feb 1, 2003).

42. S. V. Petersen, S. Thiel, L. Jensen, R. Steffensen, J. C. Jensenius, J. Immunol. Methods 257, 107 (Nov 1, 2001).

43. S. E. Degn et al., J. Immunol. Methods 373, 89 (Oct 28, 2011).

